# Urine proteomic signatures of histological class, activity, chronicity, and treatment response in lupus nephritis

**DOI:** 10.1101/2023.07.17.23292359

**Authors:** Andrea Fava, Jill Buyon, Laurence Magder, Jeff Hodgin, Avi Rosenberg, Dawit S. Demeke, Deepak A. Rao, Arnon Arazi, Alessandra Ida Celia, Chaim Putterman, Jennifer H. Anolik, Jennifer Barnas, Maria Dall’Era, David Wofsy, Richard Furie, Diane Kamen, Kenneth Kalunian, Judith A. James, Joel Guthridge, Mohamed G. Atta, Jose Monroy Trujillo, Derek Fine, Robert Clancy, H. Michael Belmont, Peter Izmirly, William Apruzzese, Daniel Goldman, Celine C. Berthier, Paul Hoover, Nir Hacohen, Soumya Raychaudhuri, Anne Davidson, Betty Diamond, Accelerating Medicines Partnership in RA/SLE network, Michelle Petri

## Abstract

Lupus nephritis (LN) is a pathologically heterogenous autoimmune disease linked to end-stage kidney disease and mortality. Better therapeutic strategies are needed as only 30-40% of patients completely respond to treatment. Noninvasive biomarkers of intrarenal inflammation may guide more precise approaches. Because urine collects the byproducts of kidney inflammation, we studied the urine proteomic profiles of 225 LN patients (573 samples) in the longitudinal Accelerating Medicines Partnership (AMP) in RA/SLE cohort. Urinary biomarkers of monocyte/neutrophil degranulation (i.e., PRTN3, S100A8, azurocidin, catalase, cathepsins, MMP8), macrophage activation (i.e., CD163, CD206, galectin-1), wound healing/matrix degradation (i.e., nidogen-1, decorin), and IL-16 characterized the aggressive proliferative LN classes and significantly correlated with histological activity. A decline of these biomarkers after 3 months of treatment predicted the 1-year response more robustly than proteinuria, the standard of care (AUC: CD206 0.92, EGFR 0.9, CD163 0.89, proteinuria 0.8, p<0.01). Candidate biomarkers were validated and provide new potentially treatable targets. We propose these biomarkers of intrarenal immunological activity as noninvasive tools to diagnose LN, guide treatment, and as surrogate endpoints for clinical trials. These findings provide new insights into the processes involved in LN activity. This dataset (matching other AMP omics) is a public resource to generate and test hypotheses and validate biomarkers.

## Introduction

Lupus nephritis (LN) is a leading cause of morbidity and mortality in patients with SLE resulting in end-stage kidney disease (ESKD) in 20% of cases (1), especially in ancestral minorities (2, 3). Despite optimal treatment, only 30-40% of LN patients achieve a complete renal response at 1 year (4–6). Thus, there is a pressing need to identify novel treatment strategies to prevent kidney damage and mortality.

LN diagnosis, classification, and treatment rely on kidney biopsies obtained in SLE patients with proteinuria. Kidney biopsies are necessary because proteinuria neither distinguishes treatable inflammation from chronic damage nor differentiates International Society of Nephrology (ISN) LN classes. Furthermore, proteinuria does not correlate with intrarenal inflammation and is a lagging indicator as it occurs and persist after damage has ensued. Kidney biopsies also have limitations, including procedure-related complications and sampling error, and may delay diagnosis and treatment. In addition, kidney biopsies are invasive and may be challenging to repeat in all patients with LN. A noninvasive biomarker that reflects intrarenal pathology could lead to early diagnosis and guide treatment by assessing response in real time.

Urine collects the byproducts of kidney pathology. By reflecting intrarenal processes (7, 8), urine proteomics is an ideal non-invasive tool to discover disease mechanism, identify novel therapeutic targets, and confirm noninvasive biomarkers. Previous studies explored urine proteomics in LN, but were limited by technical sensitivity, sample size, or cross-sectional design (7, 9-13). Several biomarkers have been identified in LN. However, their performance has been often measured as the ability to discriminate LN from healthy donors or proteinuric LN from LN in remission (14). Such biomarkers provide limited clinical value because they do not outperform readily available biomarkers (i.e. proteinuria) and thus do not impact treatment decisions or prognosis.

Accordingly, this study applied urine proteomics to the large and ancestrally diverse Accelerating Medicines Partnership LN longitudinal cohort (15) to define pathways and clinically meaningful biomarkers linked to histology class, LN activity and response to treatment. We found that markers of neutrophil/monocyte degranulation, macrophage activation, and extracellular matrix remodeling are implicated in proliferative LN (the most aggressive type), LN activity, and response to treatment. Candidate biomarkers were validated and provide new potentially treatable targets. This large dataset is available to the public for further research.

## Results

### Pipeline and recruitment

To characterize LN molecular signatures of specific LN subtypes and treatment response, we analyzed the longitudinal urine proteomic profiles (1200 proteins) of LN patients and their clinical and histologic associations (**Figure 1**).

**Figure 1.**
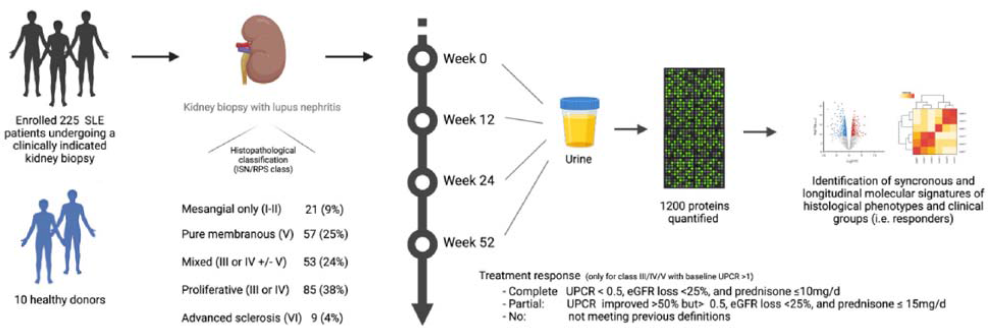
Experimental pipeline.

We recruited 225 SLE patients who underwent a clinically indicated kidney biopsy and had a urine protein to creatinine ratio > 0.5g/g. To capture LN diversity, we included all patients with LN defined by histology. Most patients (62%) had proliferative LN: 85 (38%) with pure proliferative LN (class III or IV) and 53 (24%) with mixed LN (class III or IV + V); 25% had pure membranous LN (class V); 21 (9%) had mesangial limited LN (class I or II); and 9 (4%) had advanced sclerosis (class VI). For comparison, we recruited 10 healthy donors (HD) without a past medical history of any kidney disease and negative autoimmune serologies. All patients and controls had a baseline sample. Longitudinal urine samples were collected for patients with class III, IV, or V LN; 136, 109, and 96 samples were analyzed at week 12, 24, and 52, respectively. The baseline demographic and clinical characteristics are summarized by **Table S1**. The patients were similar in age and sex. As expected, proliferative LN had higher histological activity (NIH Activity Index). Except for ISN class VI (advanced sclerosis), chronicity was similar in the other classes. Proteinuria at the time of biopsy was lower in class I or II LN (median 0.76 [range 0.5-4]) whereas all other classes were similar, highlighting the inability of proteinuria to distinguish between LN classes as we have previously shown(16). The estimated GFR was reduced in all LN patients compared to HD with the lowest values observed in class VI (median 46 ml/min [range 9-63]), followed by proliferative LN (median 88 ml/min [range 12-160]), and pure membranous (median 100 ml/min [range 15-145]). Patients with proliferative or membranous LN were followed longitudinally: complete response rates at week 52 were more common in proliferative LN as compared to pure membranous LN (34% vs 16%). We assayed a total of 573 urine proteomic profiles from these 225 unique LN patients and 10 HD.

### Molecular signatures of lupus nephritis

To identify the proteomic signature of each LN class, we initially compared the urine proteomic profile of LN with HD without clinical proteinuria (**Figure 2A-C**). Hundreds of proteins were significantly increased in all LN classes *(**Supplementary File 1**)*. In this study, pure proliferative LN (class III or IV) and mixed LN (class III or IV + V) were often analyzed together because they share the component of “proliferative” LN which is linked to worse outcomes(17). Accordingly, patients with pure proliferative and mixed LN showed similar proteomic profiles with pathway enrichment analysis detecting innate immune system, neutrophil degranulation, viral life cycle, and extracellular matrix disassembly/protease activity (**Figure 2D-H**, network analysis in **Figure S1**). Most of the proteins enriched in pure membranous LN were also found in proliferative LN, indicating common core pathways across proliferative, mixed, and pure membranous LN (**Figure 2G and S2**). In contrast, most of the proteins enriched in proliferative LN (pure or mixed) were not found in membranous suggesting that distinct biological processes are restricted to proliferative LN. At the pathway enrichment level, all three LN groups showed evidence of protease activity and extracellular matrix remodeling (**Figure 2H**).

**Figure 2.**
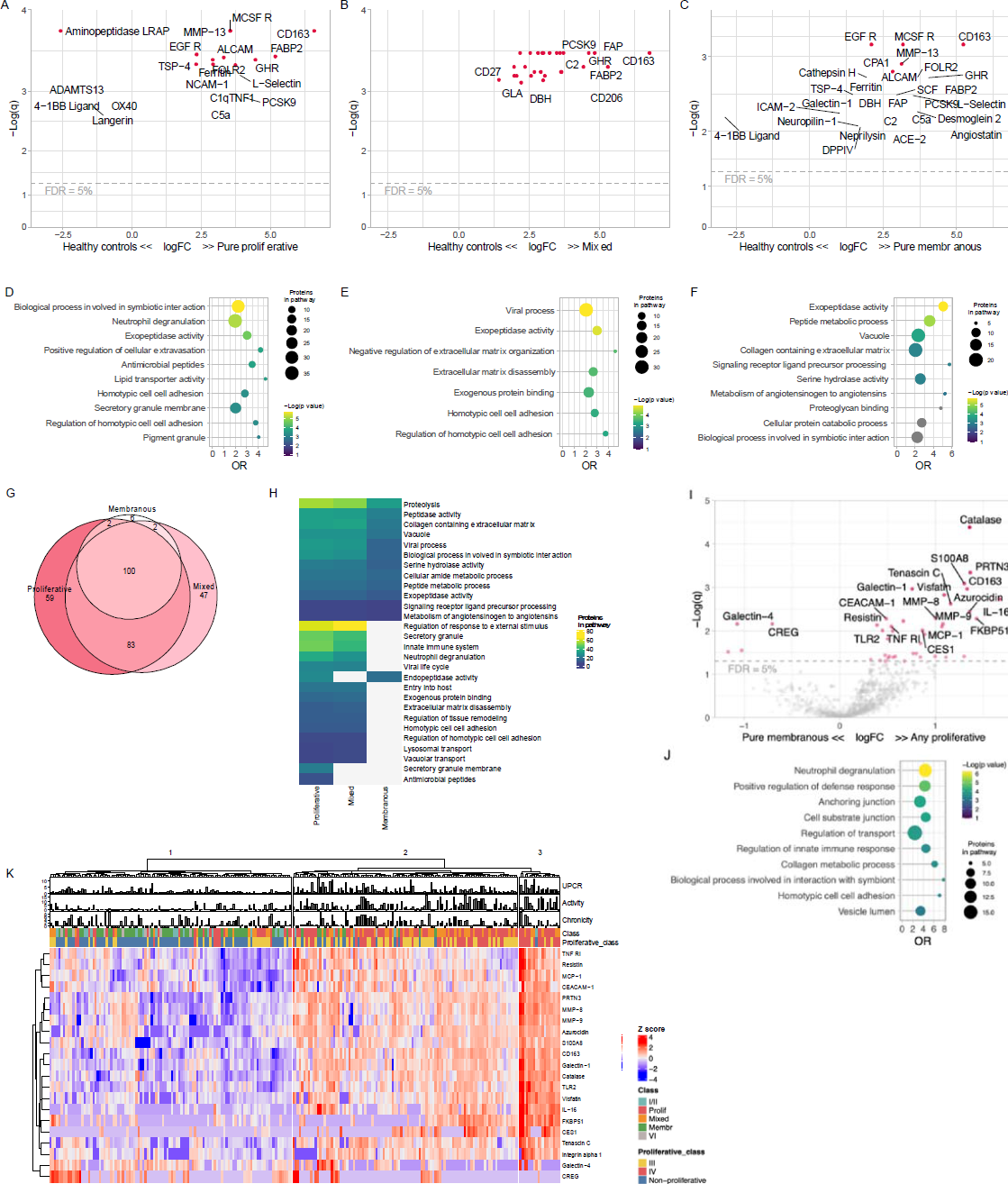
Proteomic signatures of LN histological classes. Volcano plots of the differential urinary protein abundances in pure proliferative (A), mixed (B), and membranous (C) LN compared to healthy controls (HD). Pathway enrichment analysis of the proteins enriched in pure proliferative (D), mixed (E), and membranous (F) (FDR <5%); pathways in gray had a q value > 0.05. (G) Venn diagram summarizing the shared significantly changed proteins at enriched in the 3 classes displayed in A-C. (H) Heatmap summarizing the pathways enriched (FDR <25%) in the 3 classes. (I) Volcano plot displaying the differential urine protein abundances in any proliferative (pure or mixed) and pure membranous with relative pathway enrichment analysis (J). (K) Heatmap displaying the unsupervised clustering based the urine abundances of the proteins differentially abundant in any proliferative vs pure membranous (panel I); clinical features are displayed. Not available activity and chronicity scores are indicated as −1. *FDR = false discovery rate; q = adjusted p value (Benjamini-Hochberg)*.

Because proliferative LN is characterized by an intraglomerular immune infiltrate with endocapillary hypercellularity, the identification of leukocyte mediated immunity proteomic profiles indicates that urine proteomics congruently reflected intrarenal pathology. Proliferative LN is the most aggressive form of LN and carries a higher risk of permanent kidney damage(17). To better define proliferative LN’s specific pathological pathways, we compared the proteomic profiles of proliferative (pure and mixed) to pure membranous LN. Proliferative LN signature was dominated by higher levels of CD163 (a macrophage marker; FC=2.5, q=0.001), IL-16 (a proinflammatory chemokine; FC=3.2, q=0.002), and granulocyte degranulation products such as PRTN3, S100A8, azurocidin, catalase, and MMP8 (range FC=2.5-2.6, q=4×10^−5^-5×10^−3^) among many others (**Figure 2I and S1A**). Pathway enrichment analysis confirmed that neutrophil degranulation was the biological signature most enriched in proliferative LN (**Figure 2J**). Several macrophage markers such as CD163, CD206, Galectin-1, and FOLR2 were enriched in all classes. The urinary abundance of these proteins was similar in pure and mixed proliferative LN, but higher than membranous (**Figure 2F and S2)**.

The urine abundance of the proteins differentially expressed in the proliferative LN signature is displayed in the heatmap in **Figure 2K**. We noted 3 clusters of patients defined by low, medium, or high expression of this signature. As expected, the “low” (left) cluster included almost exclusively patients with non-proliferative LN. The “medium” (center) cluster included mostly patients with proliferative LN, but also some pure class V, class I/II, and class VI LN. The “high” (right) cluster identified patients with the greatest expression of the proliferative LN signature and was comprised mostly by patients with proliferative LN. Patients in this cluster were largely class IV and demonstrated the highest activity indices. Of note, there were several patients with histologically non-proliferative LN in the “medium” cluster and 1 in the “high” cluster indicating heterogeneity in non-proliferative LN. About 18% of pure membranous LN showed strong inflammatory responses with degranulation and monocyte/macrophage activation signatures. These findings indicate a disconnect between the histological classification and the inflammatory activity detected by urine proteomics in several patients. Future adequately powered studies should explore whether urine proteomics can identify a subgroup of pure membranous patients more likely to respond to immunosuppression.

Altogether, these findings implicate active neutrophil/monocyte degranulation and macrophage activation in patients with proliferative LN. These signatures identified patients with higher activity indices. Furthermore, protease activity and extracellular matrix degradation characterized both proliferative and pure membranous LN. Importantly, these intrarenal biological processes can be noninvasively quantified in the urine.

### Proteomic signatures of histological activity and chronicity

Proliferative LN is heterogeneous in the degree of immunological activity. This is captured by the NIH Activity Index (18). High scores identify more aggressive disease associated with higher risk of kidney failure (17). Five of the six components of the NIH Activity Index (endocapillary hypercellularity, neutrophil/karyorrhexis, fibrinoid necrosis, wire loops/hyaline thrombi, and cellular/fibrocellular crescents) are exclusive to proliferative LN (class III or IV +/-V) thereby making the NIH Activity Index a quantitative measure of proliferative LN activity. To characterize the pathways and biomarkers of LN activity, we studied the correlation of the urinary proteins with the NIH Activity Index (**Figure 3A**). We found several urinary proteins directly correlated with the NIH Activity Index, topped by IL-16 and CD163 (**Figure 3A**). As supported by pathway enrichment analysis (**Figure 3B**), the signature of LN activity also included proteins associated with degranulation (i.e., PRTN3, azurocidin, visfatin, MMP8, LAMP1, catalase), macrophage activation (i.e., CD163, CD206, galectin-1), and wound healing/matrix degradation (i.e., nidogen-1, decorin) (**Figure S3**). Importantly, these associations persisted after adjusting for proteinuria or renal fibrosis (NIH Chronicity Index) in a multivariable model (**Figure S4**). These findings further support the link between proliferative LN and both myeloid cell activity / degranulation and wound healing pathways by demonstrating a direct quantitative association with proliferative LN activity, independent of proteinuria.

**Figure 3.**
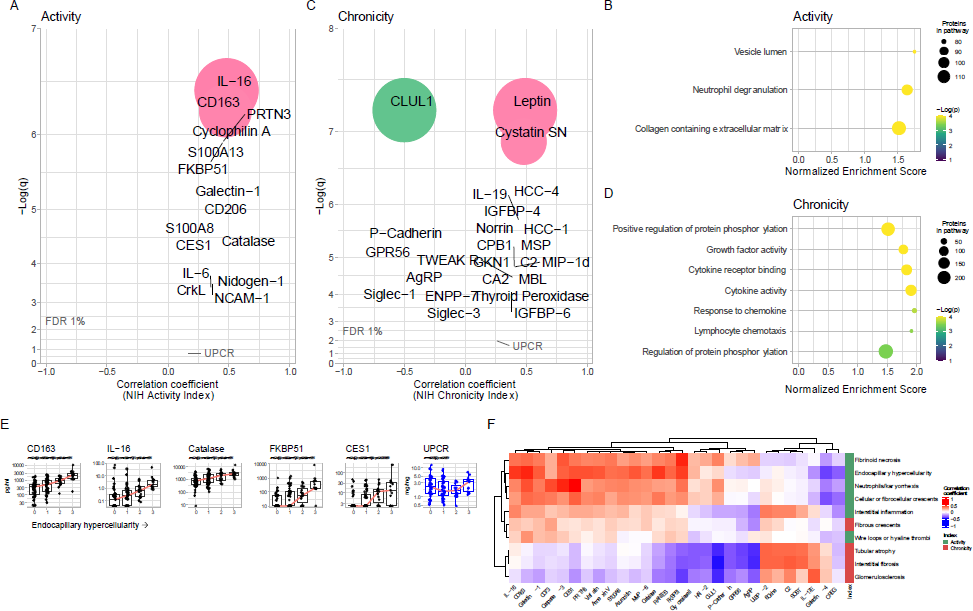
Proteomic signatures of histological activity and chronicity. Volcano plots displaying Pearson’s correlation of the proteins urinary abundances and the NIH Activity (A) and Chronicity (B) indices. The correlation with the urine protein-to-creatinine ratio (UPCR) is indicated for reference. Pathway enrichment analysis (by GSEA) of the associations of the urinary proteins with the NIH Activity (C) and Chronicity (D) indices. (E) The 5 most correlated proteins with the endocapillary hypercellularity score are displayed as compared to UPCR. (F) Hierarchical clustering based on the correlations of each histological lesion and urinary proteins. All proteins with a strict statistically significant correlation (FDR < 0.01) with at least one histological lesion were included. *FDR=false discovery rate; q=Benjamini-Hochberg adjusted p value*.

Next, we studied the proteomic correlates of intrarenal damage as quantified by the NIH Chronicity Index. The NIH Chronicity Index captures features of irreversible damage such as interstitial fibrosis and tubular damage, glomerulosclerosis, and fibrous crescents. **Figure 3C** displays the urinary proteins positively and negatively correlated with intrarenal chronicity. Pathway enrichment analysis identified cytokine/chemokines and grow factor activity (**Figure 3D**). These associations persisted after adjusting for proteinuria and the NIH Activity Index (**Figure S4**).

### Proteomic signatures of specific histological features

We analyzed the urinary proteomic profiles of each histological lesion assessed by the NIH activity and chronicity indices(18). In this sub-analysis, 115 biopsies with available subscoring were included: 42 (36%) with pure proliferative LN, 33 (29%) with pure membranous LN, and 41 (35%) with mixed LN. The five most correlated urinary proteins and each histological feature of the NIH Activity and Chronicity Index are displayed in **Figure 3E-F** and **S5**. For example, endocapillary hypercellularity, a proliferative LN-defining feature, correlated with urinary CD163, IL-16, Catalase, FKBP1, and CES1 (topping several others) but not with proteinuria (**Figure 3E**). Most lesions shared a similar signature within their respective index (**Figure 3F**). This is expected since the index components tend to co-correlate. Hierarchical clustering based on urine proteomic signatures revealed that fibrous crescents were more similar to activity-related lesions despite being considered inactive lesions and counted in the NIH Chronicity Index (**Figure 3F**). Interstitial inflammation (activity), fibrous crescents (chronicity) and, to a lesser extent, wire loops (activity) correlated with biomarkers associated with both active and chronic lesions (**Figure 3F**). Strikingly, there was no correlation between proteinuria and the histological lesions in the NIH Activity or Chronicity Indices (**Figure 3E** and **S5**).

### Treatment response is associated with a decline of urinary biomarkers of LN activity including markers of myeloid immunity and matrix degradation

Next, we focused on the proteomic signatures linked to treatment response. Complete renal response is currently defined by a decline of urine protein-to-creatinine ratio (UPCR) to < 0.5 after 1 or 2 years since it is associated with better long-term preservation of kidney function in LN. To assess response, a baseline UPCR > 1 was required (16). In this analysis, a total of 127 patients were included: 48 (38%) with pure proliferative LN (class III or IV), 41 (32%) with mixed LN (III or IV +/-V), and 38 (30%) with pure membranous LN. Response was complete in 34 (27%), partial in 29 (23%), and none in 64 (50%). In this cohort, treatment selection was at the discretion of the treating physician, but mycophenolate mofetil was the mainstay of treatment.

At the time of kidney biopsy (baseline), there was no difference in the urinary proteomic profiles in patients who achieved any clinical response at 1 year (responders) compared to non-responders (**Figure S6A**), even when the analysis was restricted to patients treated with the same regimen of mycophenolate. Therefore, we focused on longitudinal trajectories.

To identify pathways that could mediate response to immunosuppression, we studied the changes in the urinary proteome after 3 months of treatment compared to the baseline, according to the response status at 1 year. Responders showed a decline at 3 months in 69 urinary proteins (FDR <1%) led by Galectin-1, CD163, IL-16, and CD206 (**Figure 4A-B**). These proteins overlapped with the proteomic signature associated with histological activity (**Figure 3A**). Accordingly, pathway enrichment analysis after 3 months of treatment showed a decline in pathways related to extracellular matrix and cellular immune response in those who ultimately had a complete or partial response at 1 year (**Figure 4C-D**).

**Figure 4.**
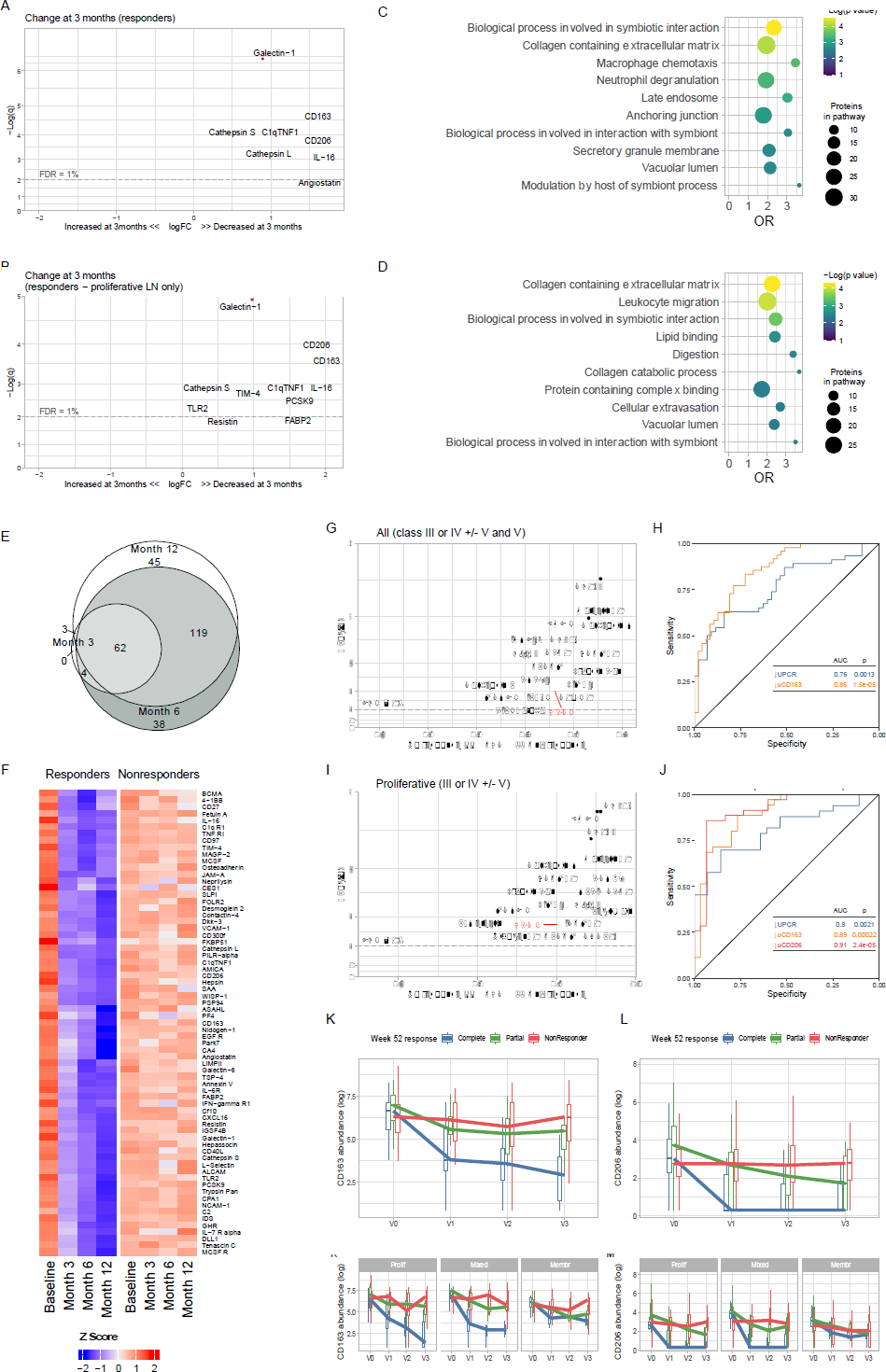
Proteomic changes of treatment response. Volcano plots of the changes of the urinary proteomic profiles of treatment responders at 3 months after kidney biopsy/treatment compared to baseline at time of biopsy in proliferative and membranous combined (A) or proliferative only (B). (C and D) Pathway enrichment analysis of the urinary proteins declined in A and B, respectively. (E) Venn diagram summarizing the shared significantly changed proteins at the 3, 6, and 12 months after the kidney biopsy. (F) Heatmap displaying the urinary abundances of the proteins significantly decreased at 3 months in responders (panel A) at the 4 time points according to response status. (G) Discriminatory power of the change of each urinary protein at 3 months compared to baseline to predict treatment response at month 12 (displayed as area under the curve, AUC). The change in urine protein-to-creatinine ratio (UPCR) is displayed for refence as the traditionally used biomarker. (H) Receiver operating characteristic curves of the decline at 3 months of the UPCR (traditional biomarker) and urinary CD163. I and J replicate G and H, but limited to patients with proliferative LN. (K) Trajectory of the urinary abundance of CD163 (K) and CD206 (L) according to response status in all patients and stratified by ISN class. Thin lines indicate individual trajectories; thick lines indicate the group medians; boxplots indicate medians, interquartile range, and range. *q values = adjusted p values (Benjamini-Hochberg). OR = odds ratio. FDR = false discovery rate.*

The decline of the urine proteins by 3 months persisted at 6 and 12 months. Moreover, an increased number of proteins declined at 6 and 12 months in responders (**Figure 4E-F and S6B-E**). By contrast, there were no changes observed in non-responders (**Figure 4F**).

To identify early biomarkers of response, we studied the discriminatory ability of the urinary protein changes at 3 months to predict 1 year response. One-hundred and eleven urinary biomarkers predicted response (FDR <1%, AUC 0.7-0.86), most outperforming the improvement in proteinuria (the clinical standard) (**Figure 4G**). A decline of CD163 at 3 months predicted 1 year response with an area under the curve (AUC) of 0.86 (q=2.7*10^−6^) compared to an AUC of 0.75 (q=0.01) for proteinuria (**Figure 4H**). In proliferative LN, urinary biomarkers displayed superior performance with an AUC of 0.91 (q=1.6*10^−5^), 0.9 (q=1.6*10^−5^), 0.89 (q=3.5*10^−5^), and 0.76 (q=0.007) for the decline of CD206, EGFR, CD163, and proteinuria, respectively (**Figure 4I-J**). In pure membranous LN, Smad4 and LAMA4 displayed AUCs of 0.88 and 0.75, respectively, with nominal p values < 0.01 but q values > 0.7. There were no biomarker changes at 3 months predicting response in pure membranous LN that reached statistical significance after correcting for multiple comparisons.

These findings indicate that effective immunosuppression induces an immunological response in the kidney by 3 months that can be noninvasively monitored in the urine. Because proliferative LN is characterized by the infiltration of intraglomerular myeloid immune cells, a decline in urinary biomarkers of myeloid inflammation in responders suggests a parallel resolution of intraglomerular inflammation. This was specific to proliferative LN as exemplified by the trajectories of CD163 and CD206 (**Figure 4K-L**).

## Discussion

The discovery of disease mechanisms, patient subgroups, biomarkers, and novel targets can be simultaneously derived from the analysis of careful phenotypes, longitudinal trajectories, and differential outcomes associated with specific interventions(19). Here, we leveraged urine proteomics to discover 1) LN biology and pathways of LN activity, and 2) biomarkers of disease activity and treatment response. We defined that neutrophil/monocyte degranulation, macrophage activation, and extracellular matrix degradation are implicated in LN activity. These processes can be noninvasively quantified and monitored in urine. Reduction of the signatures of these processes at 3 months predicted treatment response. These noninvasive urine biomarkers (such as CD163 and CD206) that parallel intrarenal inflammation outperformed the current clinical standard (proteinuria). Furthermore, this study validated IL-16 as the urinary biomarker most correlated with LN activity(8) supporting its role as a novel therapeutic target and biomarker.

### LN biology and pathways of activity

Protease activity and extracellular matrix remodeling were shared by pure membranous and proliferative classes indicating that even the less inflammatory class V LN undergoes kidney remodeling. Proliferative classes were characterized by stronger macrophage and degranulation signatures that correlated with histological activity. Macrophages are the dominant immune cell type in LN(20). CD163 (hemoglobin receptor) and CD206 (mannose receptor) exist in soluble forms as they are shed during inflammation(21). In our analysis, urinary CD163 and CD206 were increased in all classes (but at higher levels in proliferative LN), they correlated with the NIH Activity Index, and their decline best predicted treatment response. Similarly, the intrarenal abundance of CD163+ and CD206+ macrophages correlated with LN histopathological indices of LN activity (20, 22). These findings (1) confirm the association between injury-associated macrophages and LN activity and (2) indicate that the disappearance of these macrophages or their differentiation to a different phenotype anticipates better outcomes.

Several neutrophil/monocyte granule proteins (i.e., PR3 and azurocidin) in the urine were linked to LN activity implicating degranulation in proliferative LN. Azurophil granules characterize neutrophils and monocytes(23). Neutrophils, especially the subset of low-density granulocytes, have been widely implicated in SLE pathogenesis and LN (24–26). Intraglomerular neutrophils and karyorrhectic debris from apoptotic neutrophils are in fact a feature of proliferative LN and are scored in the NIH Activity Index (18). However, mature neutrophils with classical polylobate nuclei are not a dominant cell type observed in LN kidney biopsies. Rather, immature forms of neutrophils implicated in the pathogenesis of LN (27, 28) do not have polylobate nuclei suggesting that their presence in LN kidney may not be noted with traditional light microscopy(28). These less mature forms of granulocytes have enhanced ability to degranulate (28) suggesting an active role in LN. PR3 can in fact lead to extracellular matrix degradation which, in turn, can lead to fibrosis and irreversible kidney damage (29–31). Our study demonstrates the ability of urine proteomics to explore a wide array of pathological processes, including neutrophil biology which can be missed in cellular studies involving sample freezing (32). Further studies are needed to define the main cell type responsible for degranulation in LN (neutrophils, monocytes, or other myeloid cells); but also to discover if this urinary signature reflects intrarenal degranulation or spillage of circulating granule proteins.

We have previously discovered that urinary IL-16 is the protein most correlated with the NIH Activity Index (8). Here, we validated this finding, applying an unbiased approach in an independent larger cohort of LN patients, corroborating the role of IL-16 as a biomarker and in LN pathogenesis. The association of urinary IL-16 with active proliferative LN was also validated in an independent Swedish cohort (33). IL-16 is a proinflammatory chemokine that can activate and recruit CD4+ and CD9+ cells (34–37). Pro-IL-16 is cleaved into bioactive IL-16 by caspase 3(36) or PR3(38) indicating that both cell death and neutrophil/monocyte degranulation may lead to IL-16 activation. Notably, CD9 controls migration and proliferation of parietal epithelial cells in response to podocyte injury (39). CD9 stimulation mediated glomerular crescent formation and glomerular demolition(39), thereby linking IL-16 to a non-immune mechanism of proliferative LN associated with poor renal survival and mortality in LN(40, 41).

Collectively, the findings from this work indicate that following an inciting event such as immune complex deposition (42), the active phase of proliferative LN is characterized by degranulation, phagocytic/injury-associated macrophage activation, chemokine release, and extracellular matrix degradation. IL-16 may be playing a central role fueling inflammation by attracting more immune cells such as neutrophils/monocytes and promoting crescent formation. Neutrophil or monocyte degranulation may directly damage the glomerular endothelium (43) and remodel extracellular matrix promoting chronic kidney disease. It is unclear whether phagocytic and injury-associated macrophages play a regulatory or proinflammatory role in the initial phase of LN activity. Nevertheless, their disappearance or their differentiation to a different phenotype is associated with treatment response, suggesting that they track with the resolution of inflammation. Importantly, these pathogenic processes can be noninvasively monitored in the urine.

Despite fibrous crescents being considered inactive lesions that follow crescentic glomerulonephritis, urine proteomics revealed inflammatory activity associated with fibrous crescents. Thus, the presence of fibrous crescents in kidney biopsies may indicate ongoing potentially treatable inflammation. In fact, crescents are classified as fibrous if comprised by <25% of cells and fibrin and therefore a small inflammatory infiltrate can be part of fibrous crescents(18). Interstitial inflammation, which is linked to worse clinical outcomes(44), showed a distinct proteomic signature combining both activity and chronicity. This is likely because the current classification system does not separate interstitial inflammation occurring in areas with or without fibrosis. Interstitial fibrosis is in fact frequently infiltrated by immune cells. These results challenge the current interpretation of histological scores. A better understanding of the pathophysiology of processes including fibrous crescents and interstitial inflammation is needed to tailor treatment of these pathways leading to chronic damage.

### Biomarkers

This work demonstrated the feasibility of urine biomarkers to noninvasively predict clinically meaningful outcomes. Previous unbiased studies focused on the identification of biomarkers to diagnose LN vs no LN(13, 14, 45, 46). However, LN presence and activity were defined by proteinuria, a readily available biomarker. Therefore, the clinical value of the novel biomarkers over proteinuria could not be established. Other studies identified urinary biomarkers of histological activity or evaluated their longitudinal changes with treatment(14, 45, 46), but these were limited to a few selected candidate biomarkers. Here, 1) we described biomarkers to identify LN histological class and activity in lupus patients with proteinuria, the group in which renal biopsies are sought for diagnosis, and 2) we systematically studied the trajectories of 1200 potential biomarkers in relation to clinical response. These findings have important diagnostic implications.

The current classification and treatment of LN rely on histological features at the time of biopsy. A higher NIH Activity Index usually triggers more aggressive immunosuppression. In contrast, when there is a low NIH Activity Index in the presence of a high NIH Chronicity Index, proteinuria is considered secondary to damage and, therefore, not requiring new or increased immunosuppression. The unbiased catalogue of urinary biomarkers of LN (outperforming proteinuria) in this large cohort of proteinuric patients provides the basis for clinically useful biomarkers that would impact clinical decisions.

Prediction of treatment response is key to improve treatment strategies. Although there were no biomarkers at baseline that predicted response, the decline of several urinary biomarkers after 3 months of treatment strongly predicted response at 1 year. These findings underscore the power of individual trajectories to discover disease biology and to identify clinically meaningful patient subsets. Persistent elevation of the NIH Activity Index in a repeat biopsy is associated with LN flares and 44% 10-year kidney survival, compared to 100% in patients with an index of 0, regardless of resolution of proteinuria (47–50). Therefore, characterization of the pathways involved in LN activity is key to the identification of new treatable targets and biomarkers to guide diagnosis and treatment. Frequent kidney biopsies are not a practical means to judge changes in activity and chronicity indices. However, our proteomic analysis offers a feasible strategy of early and frequent monitoring. Patients with higher activity had higher urinary abundance of biomarkers of inflammation (i.e., IL-16), degranulation (i.e., PRTN3, azurocidin, catalase, MMP8, LAMP1-2), macrophage activation (i.e., CD163, CD206, galectin-1, cathepsins, MIP-1b), and extracellular matrix degradation (i.e., nidogen-1, collagens, proteoglycans). A reduction in biomarkers of these processes predicted future treatment response and outperformed proteinuria. This suggests that the effective inhibition of pathogenic mechanisms by immunosuppression can be noninvasively monitored in real time. These responses are faster than the resolution of proteinuria which requires repair of the glomerular capillary wall. A biomarker panel to noninvasively assess intrarenal activity may reshape the treatment strategy of LN based on “immunological responses” (and inform clinical trial design). For example, patients with persistent urinary biomarker elevation (indicating activity regardless of improved proteinuria) would receive more potent, different, or prolonged immunosuppression or a change in approach, while those with normalized urinary biomarker levels (indicating immunologically resolved LN) could continue and eventually safely taper potentially toxic medications. These biomarkers may guide treatment selection and clinical trial design. For example, there are currently several treatment options for LN, but the choice of the best initial treatment strategy remains unclear. In patients where urine proteomics showed no reduction in these predictors of treatment response, treatment could be rapidly modified until an immunological response is achieved without waiting for improvement in proteinuria which does not track with intrarenal inflammation. Conversely, early immunological responses in the urine proteome can reassure that the current treatment is effective. Future clinical trials and longitudinal studies should address how these urinary biomarkers of intrarenal pathology can guide treatment and whether immunological responses predict long term preservation of kidney function.

Finally, this study confirmed several known biomarkers of LN. Among others, urinary CD163(8, 51, 52), MCP-1(53), Lipocalin-2(54), and ALCAM(12) were increased in proliferative LN. Of these, only CD163 and MCP-1 correlated with the NIH Activity Index. EGF-R(55) negatively correlated with the NIH Chronicity Index and positively with the NIH Activity Index.

We acknowledge several limitations. (1) Although the proteomic assay employed here allowed for the specific and highly sensitive detection of 1,200 targets, other processes might be detected using future proteome-wide broader arrays. (2) The AMP study was an observational cohort and treatment was not homogenous as in a clinical trial. There may be biomarkers at baseline or at 3 months that better predict response to specific treatments that could not be identified. Future studies involving protocolized treatment are needed to identify drug-specific response signatures(56). (3) LN activity was quantified according to the NIH Activity Index, which is more heavily weighted on glomerular than tubulointerstitial pathology. However, it should be acknowledged that tubulointerstitial disease and other histological features are also linked to kidney survival(44).

This study showed that urine proteomics is a powerful tool to discover disease processes, nominate treatable targets, and identify noninvasive biomarkers. This dataset generated by the Accelerating Medicines Partnership (AMP) is a publicly available resource for future studies. Deep phenotyping of LN by integration of multiple omics such as kidney scRNA, multiplexed histology, digitalized histology, genetics, blood studies, and other modalities (15) in matching samples will help identify novel biomarkers, LN subgroups, and treatment strategies.

## Methods

### Study approval

Human study protocols were approved by the institutional review boards (IRBs) at each participating site, and written informed consent was obtained from all participants. Patients were enrolled at Johns Hopkins University, New York University, Albert Einstein College of Medicine, University of Rochester Medical Center, Northwell Health, University of California San Francisco, Medical University of South Carolina, University of California San Diego, Cedars-Sinai Medical Center, University of Michigan, Texas University El Paso, and University of California Los Angeles. For healthy controls, IRB approval was obtained from the University of Cincinnati and Oklahoma Medical Research Foundation. After informed consent, controls were recruited at University of Cincinnati. Samples were stored by the Oklahoma Rheumatic Disease Research Cores Center and were matched for sex, race, ethnicity, and age. Subjects were screened using a questionnaire and tested negative for the following antibodies: antinuclear, double-stranded DNA, chromatin, ribosomal P, Ro, La, Smith (Sm), SmRNP, RNP, centromere B, Scl-70, and Jo-1. Samples were processed, stored, and shipped using protocols from the Accelerating Medicines Partnership in Rheumatoid Arthritis and Systemic Lupus Erythematosus (AMP RA/SLE) Network to align with the patient samples. See Supplementary Acknowledgments for a list of members of the AMP RA/SLE Network.

### Patients and samples collection

This study enrolled SLE patients with a urine protein–to-creatinine ratio (UPCR) of >0.5 who were undergoing clinically indicated renal biopsy. Only patients with a pathology report confirming LN were included in the study. Renal biopsy sections were scored by a renal pathologist at each site according to the International Society of Nephrology (ISN)/Renal Pathology Society guidelines and the National Institutes of Health (NIH) activity and chronicity indices (18). Clinical information, including serologies, were collected at the most recent visit before the biopsy. Response status at week 52 was defined in patients with a baseline UPCR >1 as follows: complete response (UPCR ≤0.5, normal serum creatinine or <25% increase from baseline if abnormal, and prednisone ≤10 mg daily), partial response (UPCR >0.5 but ≤50% of baseline value, identical serum creatinine but prednisone dose could be up to 15mg daily), or no response (UPCR >50% of baseline value, new abnormal elevation of serum creatinine or ≥25% from baseline, or prednisone >5 mg daily). Urine specimens were acquired on the day of the biopsy (before the procedure) or within 3 weeks of the kidney biopsy. Serologic features and complement levels were assessed at the clinical visit preceding the biopsy. Proteinuria was measured on or near the day of the biopsy.

### Urine Quantibody Assay

An extended version of the Kiloplex Quantibody (RayBiotech) was used to screen urine samples as previously described (7, 8). Concentration of each analyte was normalized by urine creatinine to account for urine dilution. Urine protein abundances are expressed are pg_protein_/mg_creatinine_.

### Statistical analysis

Differential protein abundance in two groups was calculated using a Wilcoxon rank test in univariate analyses. This nonparametric test allowed for robust analysis accounting for the difference in distribution, often not normal, across the 1,200 features. We observed similar performance to logistic regression (**Figure S7**). For multivariable analyses, we used linear models or generalized linear models (R lm and glm functions) after log transforming the protein abundances (models indicated on top of the figures). To account for sparsity and large variation in the dynamic ranges of the proteins, all values for each protein abundance were added the minimum measured value before log transformation. Correlations and partial correlations (R ppcor package) were calculated on log-transformed protein abundances.

Pathway enrichment analysis was performed with the clusterProfiler or fgsea R packages using the Gene Ontology and Reactome libraries. Genes coding for the measured proteins were used. Analysis was limited to gen sets with at least 5 genes represented in the universe of the 1,200 proteins measured. To account for a limited universe of proteins (not the whole coding genome), self-contained algorithms were applied. GSEA is inherently self-contained. To define the pathways enriched in a distinct group of proteins (i.e., figure 2A-D), a hypergeometric test was used. Terms with >75% proteins overlap were removed: the term with the lowest p value was retained. All statistical tests were two-sided. All analyses were performed in R version 4.1.2.

### Data and code availability

The complete datasets used in this study will be available on the Synapse platform (synapse.org) *at the time of publication*. No custom mathematical algorithm deemed central to the conclusions was generated. Analyses can be reproduced using the publicly available versions of the R packages outlined above.

## Acknowledgments

We would like to thank the patients who participated in this study, and the scientists and clinical sites in the Accelerating Medicines Partnerships in RA/SLE. We thank Felipe Andrade and René Ferretti for their critical review of the interpretation of the results. This work was supported by the AMP Rheumatoid Arthritis and Lupus Network. AMP is a public-private partnership (AbbVie Inc., Arthritis Foundation, Bristol-Myers Squibb Co., GlaxoSmithKline plc, Lupus Foundation of America, Lupus Research Alliance, Janssen Pharmaceutica, Merck Sharp & Dohme Corp., National Institute of Allergy and Infectious Diseases, National Institute of Arthritis and Musculoskeletal and Skin Diseases, Pfizer Inc., Rheumatology Research Foundation, Sanofi, and Takeda Pharmaceuticals International Inc.) created to develop new ways of identifying and validating promising biological targets for diagnostics and drug development. See Supplemental Acknowledgments for details on network authors. This work was supported by NIH grants UH2-AR067676, UH2-AR067677, UH2-AR067679, UH2-AR067681, UH2-AR067685, UH2-AR067688, UH2-AR067689, UH2-AR067690, UH2-AR067691, UH2-AR067694, UM2-AR067678, and AR074096. The Oklahoma Rheumatic Disease Research Cores Center is funded by NIH P30AR073750. The Hopkins Lupus Cohort is funded by NIH AR 69572. Dr Fava is supported by the Jerome L. Greene Foundation, the Cupid Foundation, and the Lupus Foundation of America G-2104-01274.

## References

1. Mahajan A, Amelio J, Gairy K, Kaur G, Levy RA, Roth D, et al. Systemic lupus erythematosus, lupus nephritis and end-stage renal disease: a pragmatic review mapping disease severity and progression. Lupus. 2020;29(9):1011–20.

2. Korbet SM, Schwartz MM, Evans J, Lewis EJ, and Collaborative Study G. Severe lupus nephritis: racial differences in presentation and outcome. J Am Soc Nephrol. 2007;18(1):244–54.

3. Costenbader KH, Desai A, Alarcon GS, Hiraki LT, Shaykevich T, Brookhart MA, et al. Trends in the incidence, demographics, and outcomes of end-stage renal disease due to lupus nephritis in the US from 1995 to 2006. Arthritis Rheum. 2011;63(6):1681–8.

4. Rovin BH, Teng YKO, Ginzler EM, Arriens C, Caster DJ, Romero-Diaz J, et al. Efficacy and safety of voclosporin versus placebo for lupus nephritis (AURORA 1): a double-blind, randomised, multicentre, placebo-controlled, phase 3 trial. The Lancet. 2021;397(10289):2070–80.

5. Furie R, Rovin BH, Houssiau F, Malvar A, Teng YKO, Contreras G, et al. Two-Year, Randomized, Controlled Trial of Belimumab in Lupus Nephritis. N Engl J Med. 2020;383(12):1117–28.

6. Izmirly P, Dall’Era M, Kalunian K, K D, Kim M, Carlucci P, et al. Accelerating Medicines Partnership in SLE Network T, Petri M, Buyon J, Furie R. Longitudinal Patterns of Response to Standard of Care Therapy for Lupus Nephritis: Data from the Accelerating Medicines Partnership Lupus Network [abstract]. Arthritis Rheumatology. 2021;73.

7. Fava A, Buyon J, Mohan C, Zhang T, Belmont HM, Izmirly P, et al. Integrated urine proteomics and renal single-cell genomics identify an IFN-gamma response gradient in lupus nephritis. JCI Insight. 2020;5(12).

8. Fava A, Rao DA, Mohan C, Zhang T, Rosenberg A, Fenaroli P, et al. Urine Proteomics and Renal Single-Cell Transcriptomics Implicate Interleukin-16 in Lupus Nephritis. Arthritis Rheumatol. 2022;74(5):829–39.

9. Pejchinovski M, Siwy J, Mullen W, Mischak H, Petri MA, Burkly LC, et al. Urine peptidomic biomarkers for diagnosis of patients with systematic lupus erythematosus. Lupus. 2018;27(1):6–16.

10. Nicolaou O, Kousios A, Hadjisavvas A, Lauwerys B, Sokratous K, and Kyriacou K. Biomarkers of systemic lupus erythematosus identified using mass spectrometry-based proteomics: a systematic review. J Cell Mol Med. 2017;21(5):993–1012.

11. Zhang X, Jin M, Wu H, Nadasdy T, Nadasdy G, Harris N, et al. Biomarkers of lupus nephritis determined by serial urine proteomics. Kidney Int. 2008;74(6):799–807.

12. Stanley S, Vanarsa K, Soliman S, Habazi D, Pedroza C, Gidley G, et al. Comprehensive aptamer-based screening identifies a spectrum of urinary biomarkers of lupus nephritis across ethnicities. Nat Commun. 2020;11(1):2197.

13. Vanarsa K, Soomro S, Zhang T, Strachan B, Pedroza C, Nidhi M, et al. Quantitative planar array screen of 1000 proteins uncovers novel urinary protein biomarkers of lupus nephritis. Ann Rheum Dis. 2020;79(10):1349–61.

14. Lindblom J, Mohan C, and Parodis I. Diagnostic, predictive and prognostic biomarkers in systemic lupus erythematosus: current insights. Curr Opin Rheumatol. 2022;34(2):139–49.

15. Fava A, Raychaudhuri S, and Rao DA. The Power of Systems Biology: Insights on Lupus Nephritis from the Accelerating Medicines Partnership. Rheum Dis Clin North Am. 2021;47(3):335–50.

16. Carlucci PM, Li J, Fava A, Deonaraine KK, Wofsy D, James JA, et al. High incidence of proliferative and membranous nephritis in SLE patients with low proteinuria in the Accelerating Medicines Partnership. Rheumatology (Oxford). 2022.

17. Austin HA, 3rd, Muenz LR, Joyce KM, Antonovych TT, and Balow JE. Diffuse proliferative lupus nephritis: identification of specific pathologic features affecting renal outcome. Kidney Int. 1984;25(4):689–95.

18. Bajema IM, Wilhelmus S, Alpers CE, Bruijn JA, Colvin RB, Cook HT, et al. Revision of the International Society of Nephrology/Renal Pathology Society classification for lupus nephritis: clarification of definitions, and modified National Institutes of Health activity and chronicity indices. Kidney Int. 2018;93(4):789–96.

19. Rosen A, and Zeger SL. Precision medicine: discovering clinically relevant and mechanistically anchored disease subgroups at scale. J Clin Invest. 2019;129(3):944–5.

20. Olmes G, Buttner-Herold M, Ferrazzi F, Distel L, Amann K, and Daniel C. CD163+ M2c-like macrophages predominate in renal biopsies from patients with lupus nephritis. Arthritis Res Ther. 2016;18:90.

21. Nielsen MC, Andersen MN, Rittig N, Rodgaard-Hansen S, Gronbaek H, Moestrup SK, et al. The macrophage-related biomarkers sCD163 and sCD206 are released by different shedding mechanisms. J Leukoc Biol. 2019;106(5):1129–38.

22. Li J, Yu YF, Liu CH, and Wang CM. Significance of M2 macrophages in glomerulonephritis with crescents. Pathol Res Pract. 2017;213(9):1215–20.

23. Nichols BA, Bainton DF, and Farquhar MG. Differentiation of monocytes. Origin, nature, and fate of their azurophil granules. J Cell Biol. 1971;50(2):498–515.

24. Banchereau R, Hong S, Cantarel B, Baldwin N, Baisch J, Edens M, et al. Personalized Immunomonitoring Uncovers Molecular Networks that Stratify Lupus Patients. Cell. 2016;165(3):551–65.

25. Toro-Dominguez D, Martorell-Marugan J, Goldman D, Petri M, Carmona-Saez P, and Alarcon-Riquelme ME. Stratification of Systemic Lupus Erythematosus Patients Into Three Groups of Disease Activity Progression According to Longitudinal Gene Expression. Arthritis Rheumatol. 2018;70(12):2025–35.

26. Panousis NI, Bertsias GK, Ongen H, Gergianaki I, Tektonidou MG, Trachana M, et al. Combined genetic and transcriptome analysis of patients with SLE: distinct, targetable signatures for susceptibility and severity. Ann Rheum Dis. 2019;78(8):1079–89.

27. Evrard M, Kwok IWH, Chong SZ, Teng KWW, Becht E, Chen J, et al. Developmental Analysis of Bone Marrow Neutrophils Reveals Populations Specialized in Expansion, Trafficking, and Effector Functions. Immunity. 2018;48(2):364–79 e8.

28. Mistry P, Nakabo S, O’Neil L, Goel RR, Jiang K, Carmona-Rivera C, et al. Transcriptomic, epigenetic, and functional analyses implicate neutrophil diversity in the pathogenesis of systemic lupus erythematosus. Proc Natl Acad Sci U S A. 2019;116(50):25222–8.

29. Pezzato E, Dona M, Sartor L, Dell’Aica I, Benelli R, Albini A, et al. Proteinase-3 directly activates MMP-2 and degrades gelatin and Matrigel; differential inhibition by (-)epigallocatechin-3-gallate. J Leukoc Biol. 2003;74(1):88–94.

30. Davies M, Barrett AJ, Travis J, Sanders E, and Coles GA. The degradation of human glomerular basement membrane with purified lysosomal proteinases: evidence for the pathogenic role of the polymorphonuclear leucocyte in glomerulonephritis. Clin Sci Mol Med. 1978;54(3):233–40.

31. Turck J, Pollock AS, Lee LK, Marti HP, and Lovett DH. Matrix metalloproteinase 2 (gelatinase A) regulates glomerular mesangial cell proliferation and differentiation. J Biol Chem. 1996;271(25):15074–83.

32. Arazi A, Rao DA, Berthier CC, Davidson A, Liu Y, Hoover PJ, et al. The immune cell landscape in kidneys of patients with lupus nephritis. Nat Immunol. 2019;20(7):902–14.

33. Hayry A, Faustini F, Zickert A, Larsson A, Niewold TB, Svenungsson E, et al. Interleukin (IL) 16: a candidate urinary biomarker for proliferative lupus nephritis. Lupus Sci Med. 2022;9(1).

34. Glass WG, Sarisky RT, and Vecchio AM. Not-so-sweet sixteen: the role of IL-16 in infectious and immune-mediated inflammatory diseases. J Interferon Cytokine Res. 2006;26(8):511–20.

35. Roth S, Agthe M, Eickhoff S, Moller S, Karsten CM, Borregaard N, et al. Secondary necrotic neutrophils release interleukin-16C and macrophage migration inhibitory factor from stores in the cytosol. Cell Death Discov. 2015;1:15056.

36. Zhang Y, Center DM, Wu DM, Cruikshank WW, Yuan J, Andrews DW, et al. Processing and activation of pro-interleukin-16 by caspase-3. J Biol Chem. 1998;273(2):1144–9.

37. Qi JC, Wang J, Mandadi S, Tanaka K, Roufogalis BD, Madigan MC, et al. Human and mouse mast cells use the tetraspanin CD9 as an alternate interleukin-16 receptor. Blood. 2006;107(1):135–42.

38. Kerstein-Staehle A, Alarcin C, Luo J, Riemekasten G, Lamprecht P, and Müller A. Op0054 New Role for Proteinase 3 in Il-16 Bioactivity Control in Granulomatosis with Polyangiitis. Annals of the Rheumatic Diseases. 2021;80(Suppl 1):28.1-9.

39. Lazareth H, Henique C, Lenoir O, Puelles VG, Flamant M, Bollee G, et al. The tetraspanin CD9 controls migration and proliferation of parietal epithelial cells and glomerular disease progression. Nat Commun. 2019;10(1):3303.

40. Tao J, Wang H, Wang SX, Yu F, and Zhao MH. The predictive value of crescents in the disease progression of lupus nephritis based on the 2018 International Society of Nephrology/Renal Pathology Society Revision System: a large cohort study from China. Ren Fail. 2020;42(1):166–72.

41. Zhang W, Yuan M, Hong L, Zhou Q, Chen W, Yang S, et al. Clinical outcomes of lupus nephritis patients with different proportions of crescents. Lupus. 2016;25(14):1532–41.

42. Davidson A. What is damaging the kidney in lupus nephritis? Nat Rev Rheumatol. 2016;12(3):143–53.

43. Carlucci PM, Purmalek MM, Dey AK, Temesgen-Oyelakin Y, Sakhardande S, Joshi AA, et al. Neutrophil subsets and their gene signature associate with vascular inflammation and coronary atherosclerosis in lupus. JCI Insight. 2018;3(8).

44. Wilson PC, Kashgarian M, and Moeckel G. Interstitial inflammation and interstitial fibrosis and tubular atrophy predict renal survival in lupus nephritis. Clin Kidney J. 2018;11(2):207–18.

45. Birmingham DJ, Merchant M, Waikar SS, Nagaraja H, Klein JB, and Rovin BH. Biomarkers of lupus nephritis histology and flare: deciphering the relevant amidst the noise. Nephrol Dial Transplant. 2017;32(suppl_1):i71-i9.

46. Laskari K, Mavragani CP, Tzioufas AG, and Moutsopoulos HM. Mycophenolate mofetil as maintenance therapy for proliferative lupus nephritis: a long-term observational prospective study. Arthritis Res Ther. 2010;12(6):R208.

47. Alvarado A, Malvar A, Lococo B, Alberton V, Toniolo F, Nagaraja H, et al. The value of repeat kidney biopsy in quiescent Argentinian lupus nephritis patients. Lupus. 2014;23(8):840–7.

48. Alsuwaida A, Husain S, Alghonaim M, AlOudah N, Alwakeel J, ullah A, et al. Strategy for second kidney biopsy in patients with lupus nephritis. Nephrol Dial Transplant. 2012;27(4):1472–8.

49. De Rosa M, Azzato F, Toblli JE, De Rosa G, Fuentes F, Nagaraja HN, et al. A prospective observational cohort study highlights kidney biopsy findings of lupus nephritis patients in remission who flare following withdrawal of maintenance therapy. Kidney Int. 2018;94(4):788–94.

50. Malvar A, Pirruccio P, Alberton V, Lococo B, Recalde C, Fazini B, et al. Histologic versus clinical remission in proliferative lupus nephritis. Nephrol Dial Transplant. 2017;32(8):1338–44.

51. Mejia-Vilet JM, Zhang XL, Cruz C, Cano-Verduzco ML, Shapiro JP, Nagaraja HN, et al. Urinary Soluble CD163: a Novel Noninvasive Biomarker of Activity for Lupus Nephritis. J Am Soc Nephrol. 2020;31(6):1335–47.

52. Zhang T, Li H, Vanarsa K, Gidley G, Mok CC, Petri M, et al. Association of Urine sCD163 With Proliferative Lupus Nephritis, Fibrinoid Necrosis, Cellular Crescents and Intrarenal M2 Macrophages. Front Immunol. 2020;11:671.

53. Kiani AN, Johnson K, Chen C, Diehl E, Hu H, Vasudevan G, et al. Urine osteoprotegerin and monocyte chemoattractant protein-1 in lupus nephritis. J Rheumatol. 2009;36(10):2224–30.

54. Rubinstein T, Pitashny M, Levine B, Schwartz N, Schwartzman J, Weinstein E, et al. Urinary neutrophil gelatinase-associated lipocalin as a novel biomarker for disease activity in lupus nephritis. Rheumatology (Oxford). 2010;49(5):960–71.

55. Mejia-Vilet JM, Shapiro JP, Zhang XL, Cruz C, Zimmerman G, Mendez-Perez RA, et al. Association Between Urinary Epidermal Growth Factor and Renal Prognosis in Lupus Nephritis. Arthritis Rheumatol. 2021;73(2):244–54.

56. Weeding E, Fava A, Mohan C, Magder L, Goldman D, and Petri M. Urine proteomic insights from the belimumab in lupus nephritis trial. Lupus Sci Med. 2022;9(1).

