## Supplemental material for "Urine proteomic signatures of histological class, activity, chronicity, and treatment response in lupus nephritis"

Fava A, et al.

#### Supplementary Table

1. Clinical and demographic characteristics.

#### Supplementary Figures

1. Network analysis of proteomic profiles of LN classes
2. Protein profiles of LN classes.
3. Network analysis of the pathways correlating with histological activity and chronicity.
4. Multivariable analysis of proteomic signatures of histological activity and chronicity.
5. Proteomic signatures of lupus nephritis histological lesions.
6. Additional proteomics signatures associated with response
7. Models' comparison.

### Table S1

| | Overall | HD | Lupus nephritis | | | | | p |
|---|---|---|---|---|---|---|---|---|
| | | | I/II | III or IV | III or IV +/- V | V | VI | |
| n | 235 | 10 | 21 | 85 | 55 | 55 | 9 | |
| Age (mean (SD)) | 36.4 (11.8) | 38.3 (14.8) | 39.4 (11.9) | 34.6 (11.3) | 36.7 (11.7) | 36.6 (11.5) | 41.2 (13.8) | 0.394 |
| Sex, F/M (%) | 202/33 (86.0/14.0) | 8/2 (80.0/20.0) | 20/1 (95.2/4.8) | 76/9 (89.4/10.6) | 44/11 (80.0/20.0) | 46/9 (83.6/16.4) | 8/1 (88.9/11.1) | 0.472 |
| Race (%) | | | | | | | | 0.499 |
| Asian | 35 (14.9) | 1 (10.0) | 5 (23.8) | 13 (15.3) | 8 (14.5) | 6 (10.9) | 2 (22.2) | |
| Black | 99 (42.1) | 3 (30.0) | 11 (52.4) | 32 (37.6) | 18 (32.7) | 30 (54.5) | 5 (55.6) | |
| White | 72 (30.6) | 6 (60.0) | 4 (19.0) | 28 (32.9) | 20 (36.4) | 13 (23.6) | 1 (11.1) | |
| Other | 6 (2.6) | 0 (0.0) | 1 (4.8) | 3 (3.5) | 1 (1.8) | 1 (1.8) | 0 (0.0) | |
| Unknown | 23 (9.8) | 0 (0.0) | 0 (0.0) | 9 (10.6) | 8 (14.5) | 5 (9.1) | 1 (11.1) | |
| First biopsy (%) | 82 (34.9) | 0 (0.0) | 6 (28.6) | 41 (48.2) | 20 (36.4) | 15 (27.3) | 0 (0.0) | 0.002 |
| Proliferative class (%) | | | | | | | | <0.001 |
| III | 78 (33.2) | | | 46 (54.1) | 32 (58.2) | | | |
| IV | 60 (25.5) | | | 39 (45.9) | 21 (38.2) | | | |
| NIH Activity Index (median [IQR])[1] | 4 [1, 8] | | 1 [0, 2] | 5 [3, 9] | 7 [5, 11.5] | 0 [0, 1] | 0 [0, 0] | <0.001 |
| NIH Chronicity Index (median [IQR])[1] | 3 [1, 5] | | 3 [2.5, 4] | 2 [1, 4] | 3 [2, 5.25] | 3 [1, 5.75] | 9[3] | 0.113 |
| UPCR (mean (SD)) | 2.72 (2.39) | | 1.30 (1.09) | 2.39 (1.74) | 3.62 (3.23) | 2.90 (2.42) | 2.16 (1.13) | 0.002 |
| Serum creatinine mg/ml at biopsy (mean (SD)) | 1.18 (0.82) | 0.76 (0.15) | 1.00 (0.43) | 1.23 (0.91) | 1.21 (0.77) | 1.04 (0.61) | 2.25 (1.37) | 0.001 |
| eGFR ml/min at biopsy (mean (SD)) | 85.9 (35.9) | 109.1 (19.1) | 88.6 (33.6) | 84.0 (36.3) | 83.2 (35.3) | 94.2 (34.7) | 38.4 (16.7) | <0.001 |
| Low C3 (%) | 130 (60.2) | | 5 (31.2) | 65 (79.3) | 35 (64.8) | 24 (43.6) | 1 (11.1) | <0.001 |
| Low C4 (%) | 107 (49.5) | | 7 (43.8) | 55 (67.1) | 28 (51.9) | 17 (30.9) | 0 (0.0) | <0.001 |
| Response status[2] (%) | | | | | | | | 0.28 |
| Complete | 34 (14.5) | | | 16 (18.8) | 13 (23.6) | 5 (9.1) | | |
| Partial | 29 (12.3) | | | 11 (12.9) | 9 (16.4) | 9 16.4) | | |
| No | 64 (27.2) | | | 21 (24.7) | 20 (36.4) | 23 41.8) | | |

**Supplementary Table 1. Clinical and demographic characteristics.**

[1] Activity and Chronicity Indices were available for 154 patients.
[2] Response status was defined only for patients with baseline UPCR >1.
[3] Chronicity Index available for only 1 patient.
UPCR: urine protein-to-creatinine ratio.

### Figure S1

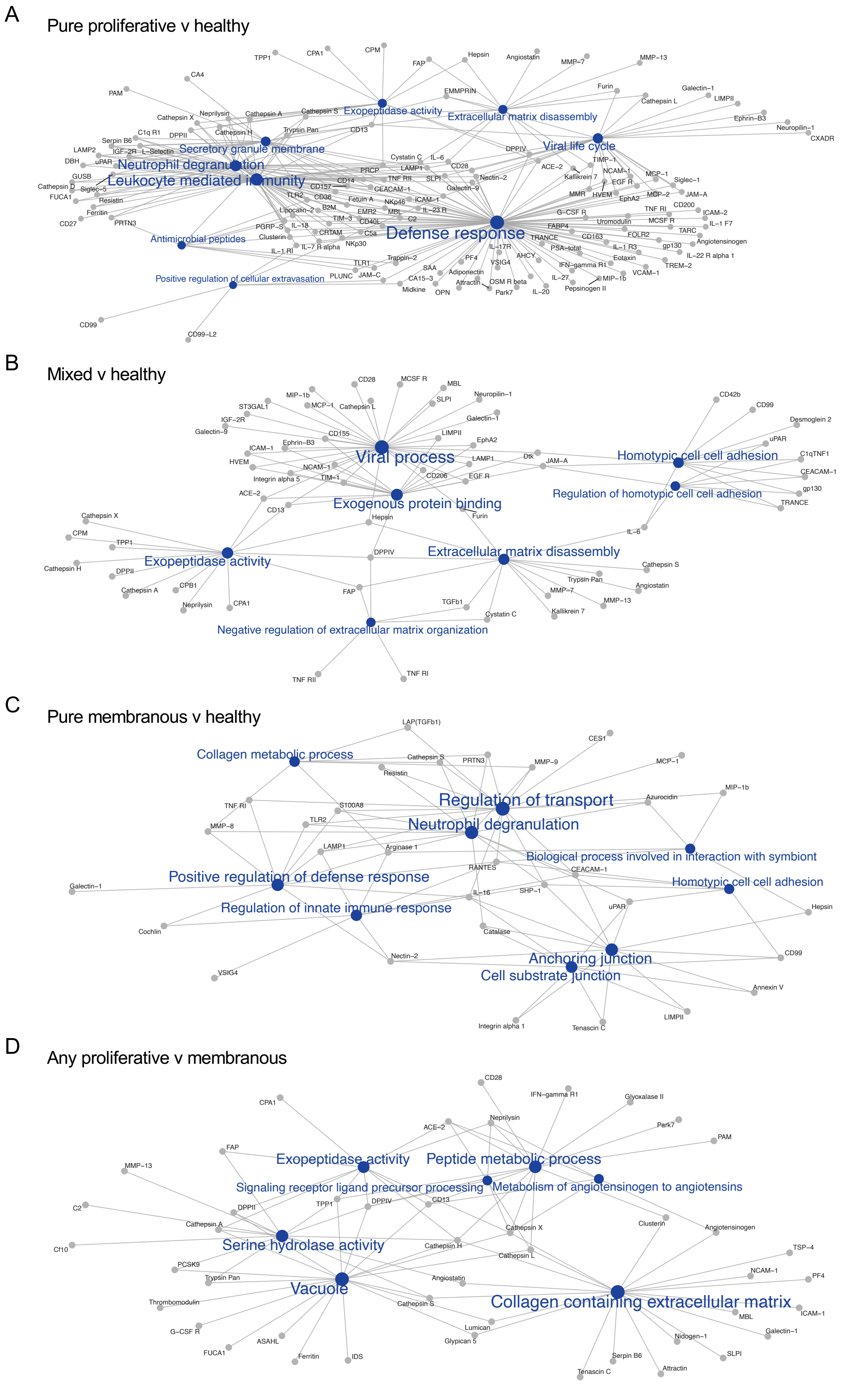


**Figure S1. Network analysis of proteomic profiles of LN classes.** Network analysis of the top 10 (or all with FDR <5%) enriched pathways comparing the urine proteomic profiles of LN classes with healthy donors or between classes.

### Figure S2

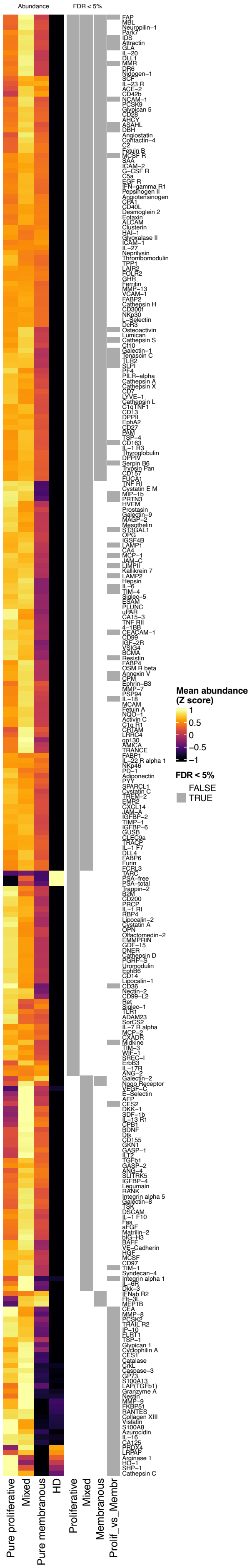


**Figure S2. Proteomic profiles of LN classes.** Heatmap displaying the urinary abundance (row normalized, Z score) of the proteins significantly enriched (FDR < 5%) in each class (fig. 2A-C) or comparing proliferative with membranous LN (fig. 2I). A gray square in the right panel indicate if the protein was significantly enriched (FDR < 5%) in the urine of the class indicated compared to healthy controls (first 3 columns) or comparing proliferative with membranous LN (last column).

**Figure S3. Network analysis of the pathways correlating with histological activity and chronicity**

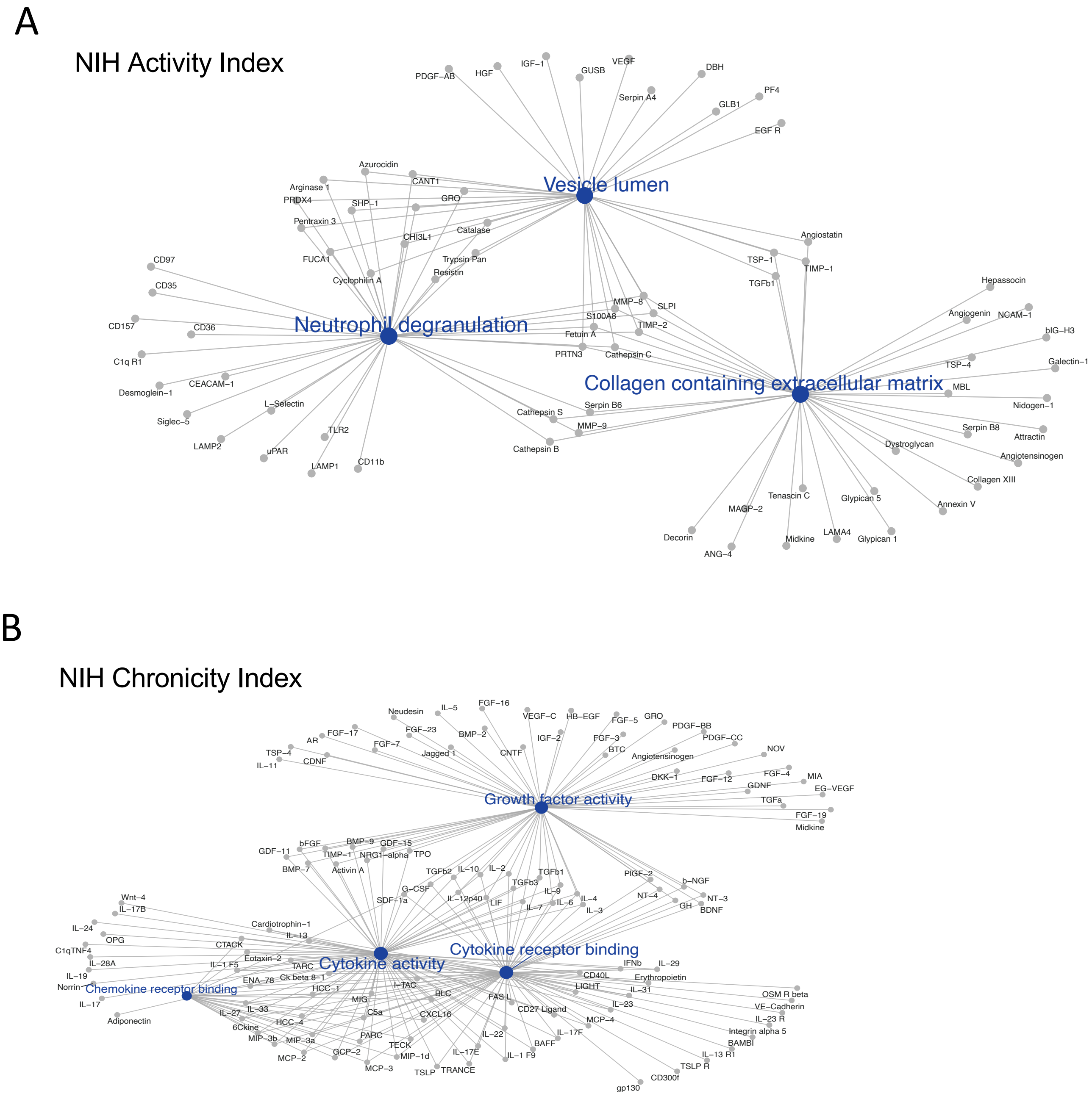


**Figure S3. Network analysis of the pathways correlating with histological activity (A) and chronicity (B).**

### Figure S4. Multivariable analysis of proteomic signatures of histological activity and chronicity.

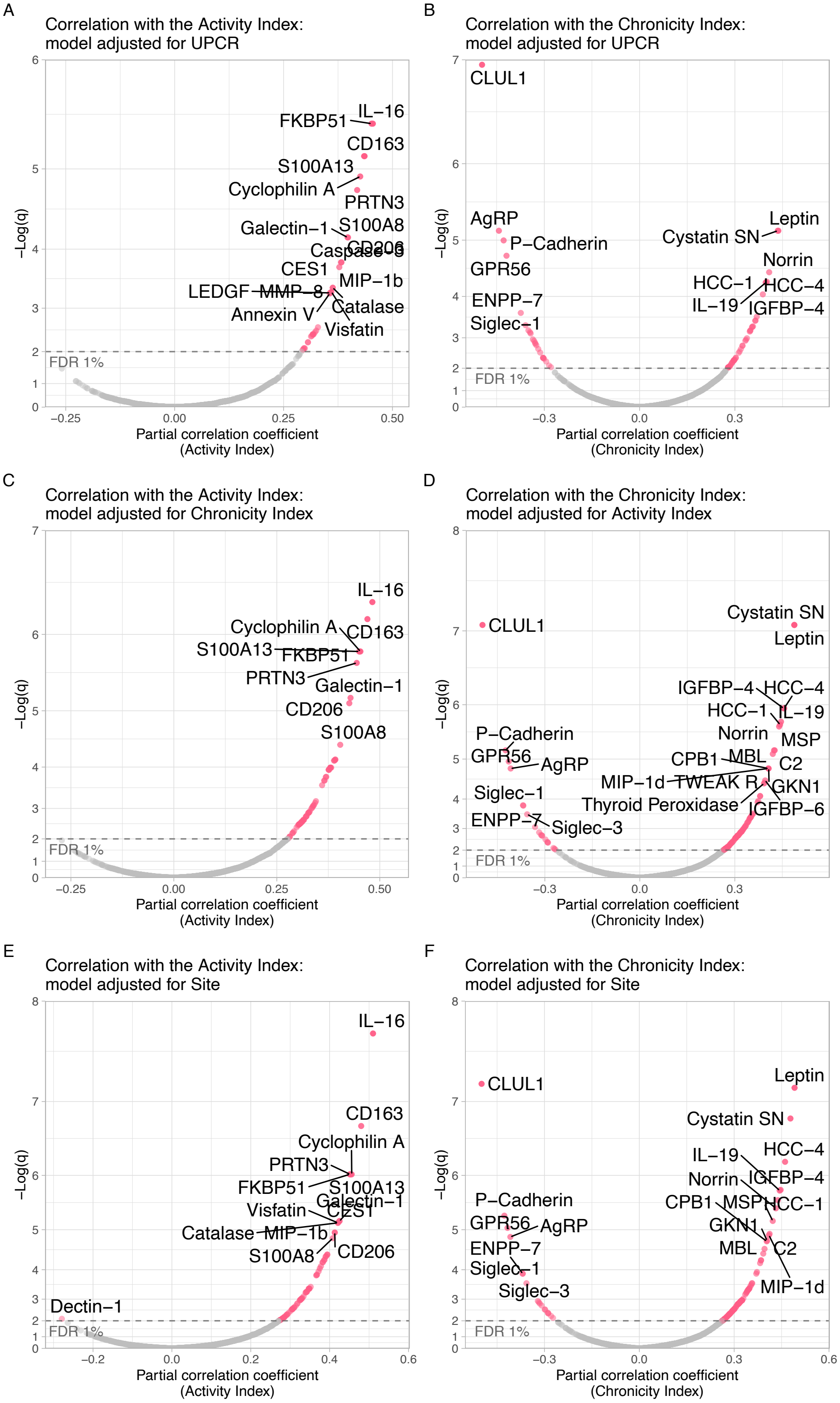


**Figure S4. Multivariable analysis of proteomic signatures of histological activity and chronicity.** Volcano plots displaying Pearson's partial correlation of the proteins urinary abundances and the NIH Activity and Chronicity indices after adjusting in a multivariable linear model for proteinuria (UPCR) (A-B), the NIH Chronicity or Activity Index (C-D), and site (E-F). *FDR=false discovery rate; q=Benjamini-Hochberg adjusted p value.*

### Figure S5. Proteomic features of lupus nephritis histological lesions

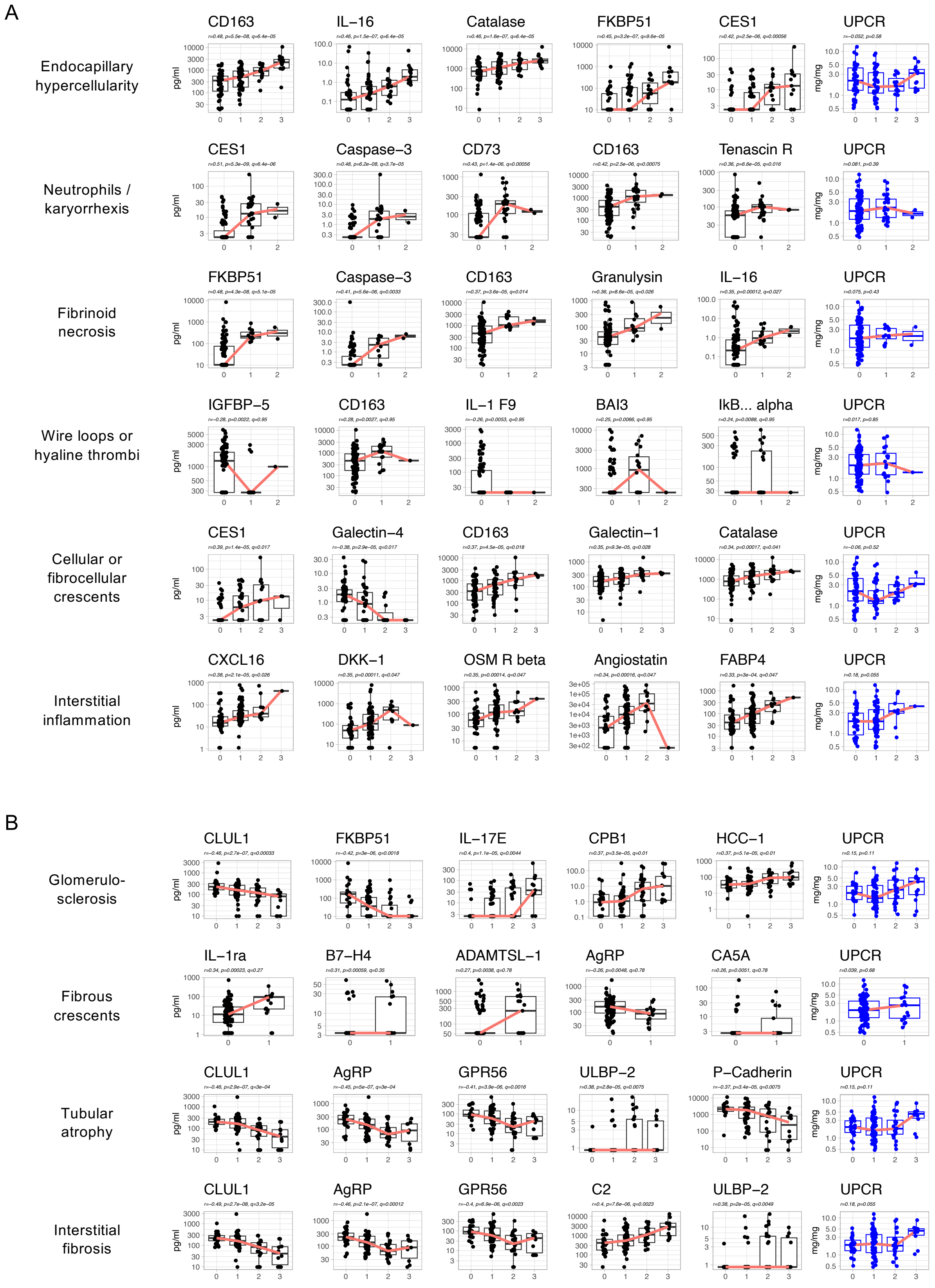


**Figure S5. Proteomic signatures of histological activity and chronicity lesions.** The 5 most correlated proteins with each histological lesion in the NIH Activity and Chronicity Indices summarized in figure 3F-G are displayed in A and B as compared to UPCR. Spearman's correlation coefficients, p , and adjusted p values (q) are shown)

**Figure S6. Additional proteomic signatures associated with response.**

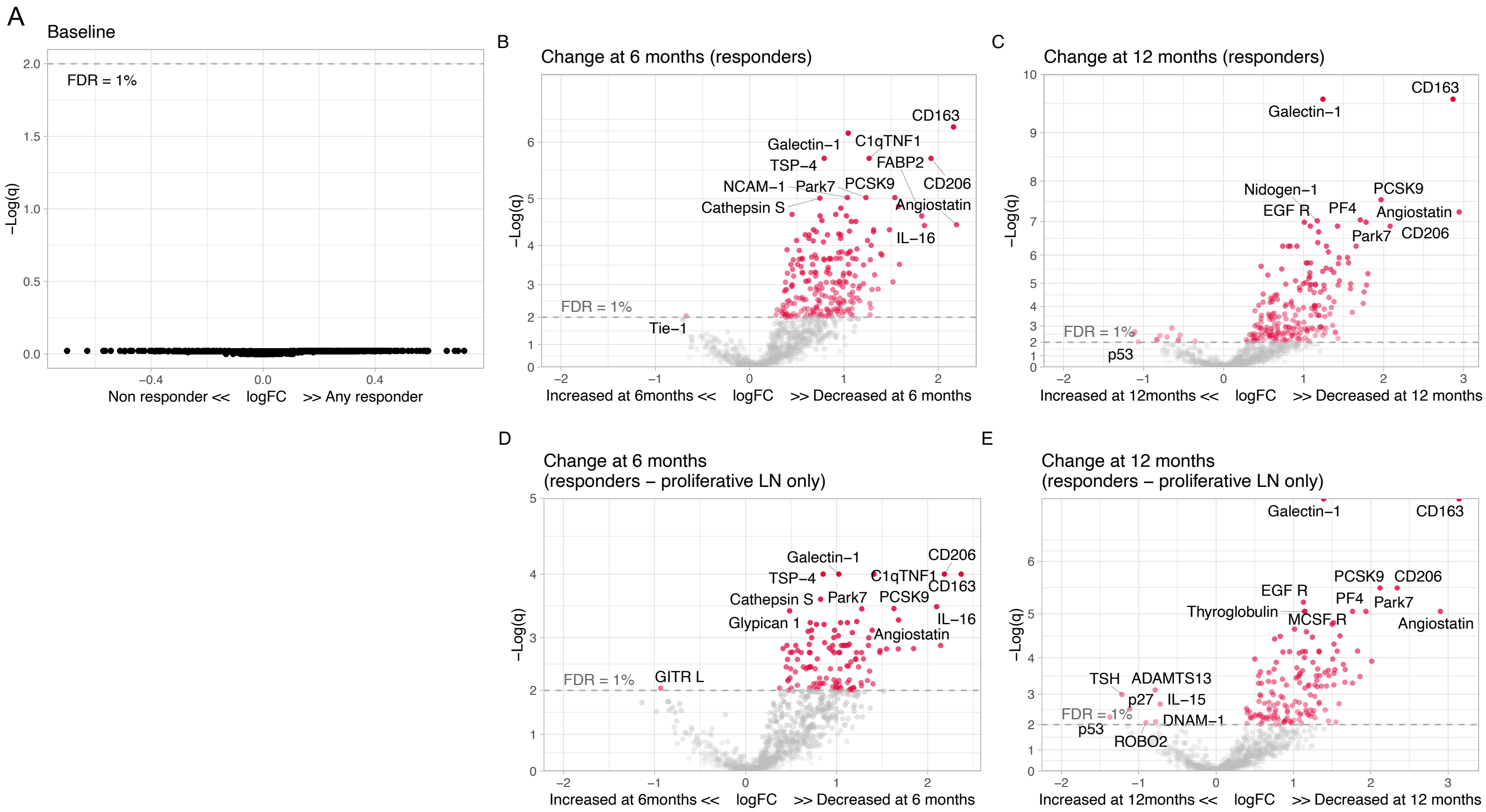


**Figure S6. Additional proteomic signatures associated with response.** (A) Volcano plots displaying that no urinary protein abundances at baseline were different in 12-month responders compared to non responders. Volcano plots of the changes of the urinary proteomic profiles of treatment responders at 6 (B) and 12 (C) months after kidney biopsy/treatment compared to baseline at time of biopsy in proliferative and membranous combined. D and E replicate panels B and C, but limited to proliferative LN. *q values = adjusted p values (Benjamini-Hochberg). FC = fold change. FDR = false discovery rate.*

**Supplementary Fig 7. Models' comparison.**

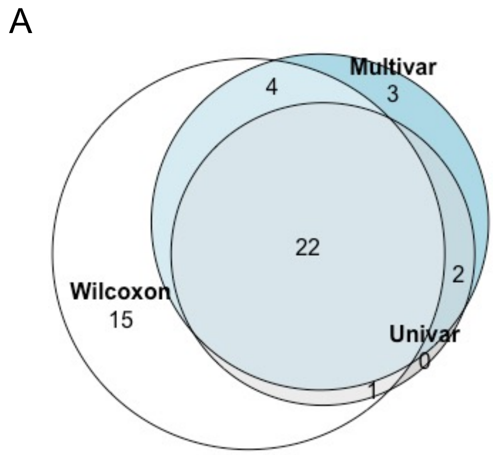


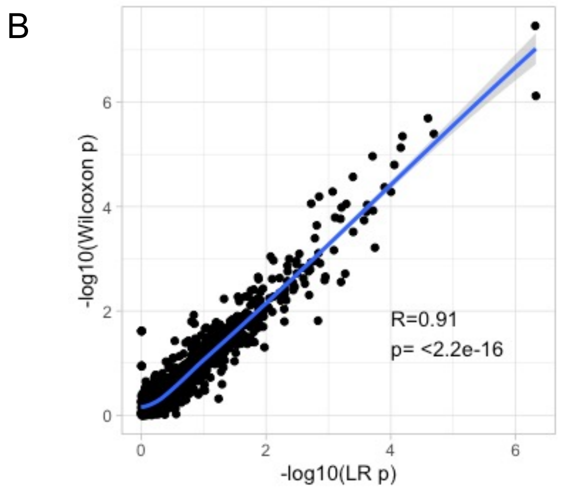


**Supplementary Fig 7. Models' comparison.** (A) Overlap of differential expressed urinary proteins between any proliferative and pure membranous LN using 3 models: Wilcoxon, univariate logistic regression (class ~ protein_abundance), and multivariate logistic regression (class ~ protein_abundance + UPCR). An FDR of <5% was considered statistically significant. (B) Correlation of p values for the association of each of the urinary protein abundance with the outcome variable (any proliferative or pure membranous LN ) using the rank-based Wilcoxon test or logistic regression (LR). Blue line displaying the loess curve. Pearson's correlation coefficient and p values are displayed.
